# Deep Learning Study of Alkaptonuria Spinal Disease Assesses Global and Regional Severity and Detects Occult Treatment Status

**DOI:** 10.1101/2025.03.11.25323762

**Authors:** Kendall A. Flaharty, Vibha Chandrasekar, Irene J. Castillo, Dat Duong, Carlos R. Ferreira, Suzanna Ledgister Hanchard, Ping Hu, Rebekah L. Waikel, Francis Rossignol, Wendy J. Introne, Benjamin D. Solomon

**Author notes:** Corresponding Author:* Kendall A. Flaharty, 10 Center Dr, Bethesda, MD 20892. These authors should be considered joint first author (Kendall A. Flaharty, Vibha Chandrasekar). These authors should be considered joint senior author (Francis Rossignol, Wendy J. Introne, Benjamin D. Solomon).

## Abstract

Deep learning (DL) is increasingly used to analyze medical imaging, but is less refined for rare conditions, which require novel pre-processing and analytical approaches. To assess DL in the context of rare diseases, we focused on alkaptonuria (AKU), a rare disorder that affects the spine and involves other sequelae; treatments include the medication nitisinone. Since assessing X-rays to determine disease severity can be a slow, manual process requiring considerable expertise, we aimed to determine whether our DL methods could accurately identify overall spine severity, severity at specific regions of the spine, and whether DL could detect whether patients were receiving nitisinone. We evaluated DL performance versus clinical experts using cervical and lumbar spine radiographs. DL models predicted global severity scores (30-point scale) within 1.72 ± 1.96 points of expert clinician scores for cervical and 2.51 ± 1.96 points for lumbar radiographs. For region-specific metrics, we assessed the degree of narrowing, calcium, and vacuum phenomena at each intervertebral space (IVS). Our model’s narrowing scores were within 0.191-0.557 points from clinician scores (6-point scale), calcium was predicted with 78–90% accuracy (present, absent, or disc fusion), while vacuum disc phenomenon predictions were less consistent (41–90%). Intriguingly, DL models predicted nitisinone treatment status with 68–77% accuracy, while expert clinicians appeared unable to discern nitisinone status (51% accuracy) (p = 2.0 × 10^-9^). This highlights the potential for DL to augment certain types of clinical assessments in rare disease, as well as identifying occult features like treatment status.

## Introduction

Deep learning (DL) is increasingly employed in medical practice and research. While DL is a powerful tool, many questions remain about applications to smaller datasets, such as relevant to rare conditions. For example, DL usually requires large amounts of training data, and for smaller datasets such as involving rare genetic conditions, it is important to understand how well DL models can be finetuned and how publicly available data from more common diseases can be leveraged.^1-4^

Other work has explored related issues; for example, in radiology, there has been considerable effort in analyzing DL performance and providing large datasets that can be applied to other analyses.^5^ While chest X-ray image datasets are readily available, other types of radiographs remain less explored. Publicly available and annotated cervical and lumbar spine x-ray datasets are relatively scarce, and only recently the CSXA and BUU-LSPINE datasets released 4963 cervical and 3600 spine lumbar radiographs with annotations for each vertebrae, respectively.^6,7^ These datasets focused on conditions involving lordosis or spondylolisthesis, which are more common than most genetic conditions affecting the spine.

In this paper, we investigated DL analyses of cervical and lumbar spine radiographs of individuals with alkaptonuria (AKU). AKU, the first human disorder described with autosomal recessive inheritance, is a rare inborn error of metabolism, with a prevalence of ∼1 in 250,000 births.^8,9^ AKU occurs due to pathogenic variants in the *HGD* gene, resulting in deficient homogentisate 1,2-dioxygenase. This enzyme is involved in degradation of homogentisic acid (HGA); enzyme deficiency results in increased HGA. Sequelae include dark urine, ochronosis, hypothyroidism, cardiovascular disease, and arthritis affecting the large joints and spine.^10^ Cervical and lumbar spine radiographs are often used to assess disease severity. Management is multi-faceted; nitsinone, which inhibits HGA production, is approved for AKU treatment in Europe, and is under investigation in the United States.^11-13^

Our motivations for the DL applications of AKU are as follows. First, expert clinicians and clinical researchers may need to manually and extensively annotate the sequelae of the spine to assess disease severity. This annotation requires expertise and time-consuming analysis. Second, when assessing a specific area like a single intervertebral space (IVS), clinicians may benefit from being able to holistically consider the overall appearance of the spine and other landmarks and features visible on a radiograph. By training on entire images, DL models may similarly incorporate this type of context in its analyses. Finally, although AKU progression is continuous, manual scoring requires human annotators to label AKU severity on a discretized scale. A DL model can be trained to produce continuous values, which may better align with disease progression.

Our classifier estimates AKU spine disease in two different ways: global severity for the entire cervical or lumbar spine and region-specific severity of each IVS. As AKU progresses, the intervertebral spaces narrow, often leading to calcification and vacuum disc phenomenon.^14^ Thus, a straightforward disease assessment is to predict a single global severity score for an entire radiograph. However, since each IVS contributes to global severity (and as it may be clinically important to consider specific parts of the spine in addition to overall severity), we also trained a multi-label classifier to estimate the region-specific severity at each IVS based on an entire radiograph.

Finally, while we initially focused on severity, we found that DL may be trained to detect whether a person is being treated with nitisinone. This aligns with previous publications in which DL models can detect findings occult to human experts.^15,16^

## Methods

AKU radiographs were obtained via IRB-approved studies at the NIH Clinical Center from January 2003 to May 2023. Of the 409 sets of lateral radiographs, we analyzed 397 cervical and 395 lumbar images in total (Figure 1, Table S1). We chose not to analyze thoracic or anteroposterior images due to incomplete data sets as well as advice from clinical experts regarding their interpretability (see Supplementary materials). Using EfficientNet as the base architecture, we trained independent models to evaluate the following: global severity, region-specific severity, and nitisinone treatment (Figure 1).^17^ See Supplementary material for more details about dataset collection and preparation, model selection, and expert radiographic scoring criteria.

**Figure 1.**
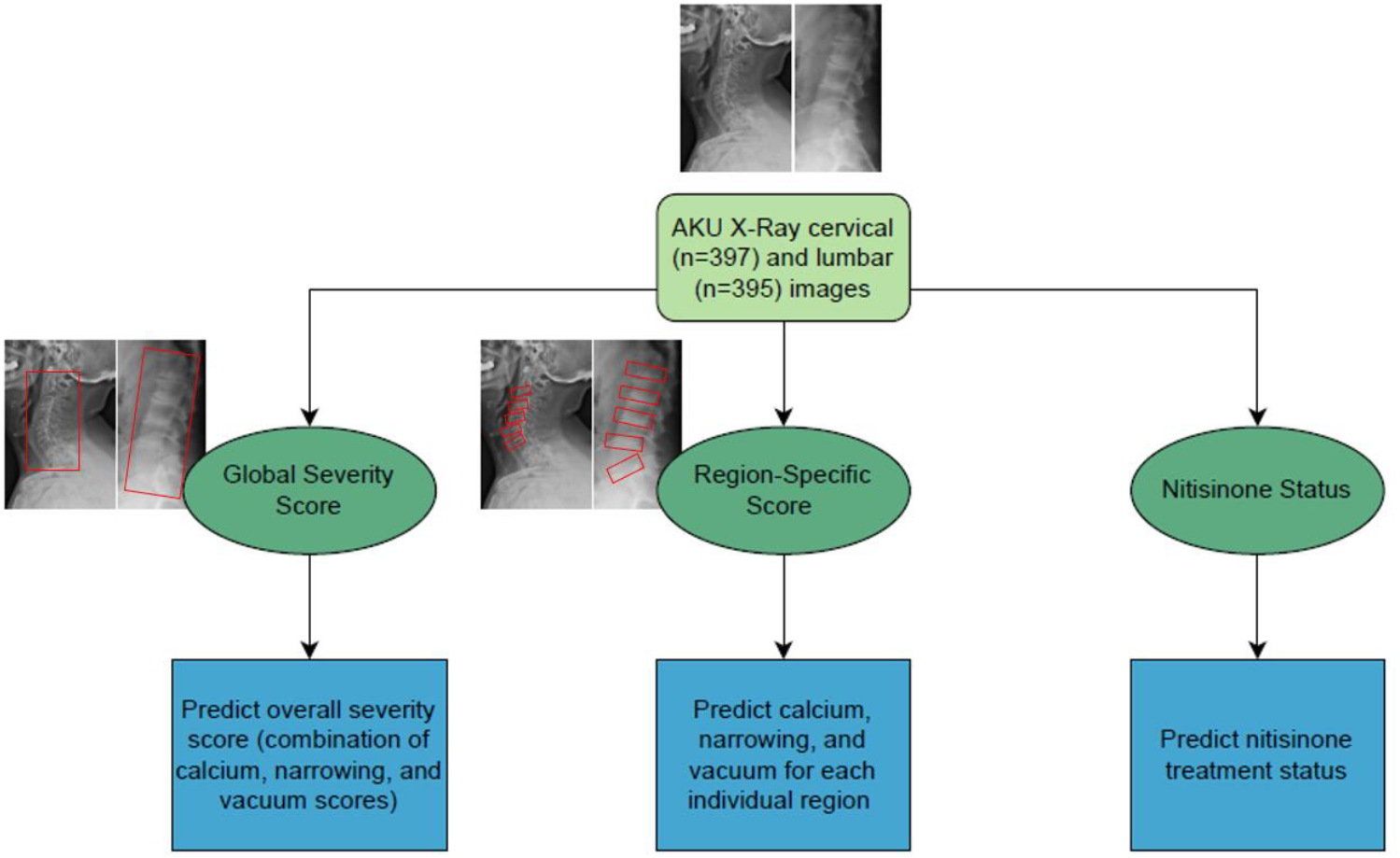
Overview of EfficientNet models trained on AKU cervical and lumbar radiographs for predicting global severity scores, region-specific scores, and nitisinone treatment status. The red boxes in the set of radiograph images indicate the specific regions of interest analyzed to attribute scores for global severity and region-specific severity, with these annotated areas serving as the input data used to train the model. One set of AKU images includes one lateral cervical and one lateral lumbar X-ray taken of a patient at the same timepoint.

### Global severity score

We estimated the global severity score of an entire radiograph, which represents a direct radiological assessment of clinical status (see Supplementary materials). The global severity scores were calculated by summing calcium (0,1), narrowing (0,1,2,3, complete disc fusion), and vacuum disc (0,1) scores across all IVS, with a maximum score of 6 per IVS (maximum 30 points per image). See Supplementary material for details about this scoring system, which was based on previous studies and per ongoing current research into spinal sequelae of AKU.^18^ A score of 6 was assigned for complete disc fusion at a given IVS; this is considered the maximum severity score for an IVS (i.e., calcium and vacuum disc were not graded for IVS with complete fusion).

We trained two classifiers, one for cervical and one for lumbar images. To mimic the continuous spectrum of AKU severity, we used soft-labels as target outputs. For each image, the ground-truth global severity score was divided by the maximum score; thus, each image has a score ranging from 0-1 instead of 0-30. For example, an image with a total score of 20 received a 20/30 soft-label target. The model was trained with Cross Entropy loss. The models were evaluated on the test set to estimate the global severity score for each radiograph and occlusion maps were generated.^19^ For human interpretation, predicted scores were rescaled from 0-1 back to 0-30 range by multiplying by 30, the maximum score. The average point differential from the ground truth score is reported for the entire test set, as well as for the lower quartile, interquartile range, and upper quartile of ground truth scores (0-25%, 25-75%, 75-100%). Linear regression R-squared values were computed to gauge how score predictions align with ground truth labels.

### Region-specific score

Next, six classifiers were trained to estimate the IVS-specific narrowing, calcification, and vacuum disc metrics for cervical and lumbar radiographs. In each classifier, every IVS was given its own Cross Entropy loss function.

Narrowing level (although continuous) was manually discretely labeled as 0, 1, 2, 3, or complete disc fusion (in which case, fusion was converted into a numerical score of 6, the maximum IVS-specific score). For prediction, we took a weighted average to estimate the final narrowing severity level in the range of 0-6.

Calcium and vacuum disc ground-truth labels were assigned by experts with discretized labeling: present, absent, or complete disc fusion (in which case, calcium and vacuum disc were not graded). For each classifier, occlusion maps were generated to assess results.^19^

### Nitisinone status

There are no obvious indicators of nitisinone treatment identifiable by human experts in radiographs; thus, as an interesting experiment, we aimed to determine if the model could differentiate treatment status.

The “on treatment” cohort included individuals who had been on nitisinone for at least three months; post-treatment individuals (i.e., individuals who had previously been treated, but who were no longer receiving treatment) were excluded. Two EfficientNet classifiers (cervical and lumbar) were trained to classify nitisinone treatment as “on” or “off”. Each model was evaluated on the same test set described in the previous sections, and occlusion maps were generated.^19^

To check whether experts could identify nitisinone status, we designed surveys using our nitisinone test images. Participants were asked whether the individual in each radiograph was receiving nitisinone (see Supplementary materials). Responses were compared to model predictions on the same test set.

We evaluated potential confounders that could be associated with nitisinone status by comparing (via *t*-test) the “on” and “off” nitisinone groups for variables such as age, time on treatment, and severity score.

## Results

### Global severity score

Table 1a presents global score predictions for cervical and lumbar radiographs. For all cervical images, the model obtained an average score 1.72 ± 1.96 points from the expert scores. For lumbar images, the range was slightly higher, with an average of 2.51 ± 1.96 points from the expert scores. Breakdowns for quartile subgroups are shown in Table 1a.

**Table 1.** DL model predictions of (a) global (total) severity scores for the full cervical and lumbar spines, (b) narrowing, calcium, and vacuum disc status for all intervertebral spaces in the cervical and lumbar spine, and (c) nitisinone status for the full cervical and lumbar spines. Global scores are reported as the average point differential with the standard deviation between the DL prediction and clinical expert annotations. Narrowing status is graded using a soft label approach, assigning levels (0, 1, 2, 3, or disc fusion, which is assigned a score of 6). Narrowing scores are reported as the average point differential with the standard deviation between the DL prediction and clinical expert annotations. Calcium and vacuum disc statuses are categorized as present, absent, or fused, and reported as the accuracy of the model in making the correct prediction. Nitisinone status is categorized as “on treatment” or “off treatment”. Individuals on the drug for less than 3 months, as well as any post-treatment images, were excluded from the cohort.

| 1a. Global Scores |  |  |  |  |
| --- | --- | --- | --- | --- |
| Region | Q1 (0-25%) | Q2-Q3 (25-75%) | Q4 (75-100%) | All Images |
| Cervical Spine | 0.72 ± 0.78 | 1.64 ± 1.44 | 3.25 ± 2.86 | 1.72 ± 1.96 |
| Lumbar Spine | 3.60 ± 2.37 | 2.18 ± 1.64 | 2.04 ± 1.56 | 2.51 ± 1.96 |
| 1b. Region-Specific Scores |  |  |  |  |
| Metric | Region | Intervertebral Space | Test Accuracy |  |
| Narrowing | Cervical Spine | C2-C3 | 0.330 ± 0.424 |  |
|  |  | C3-C4 | 0.191 ± 0.292 |  |
|  |  | C4-C5 | 0.270 ± 0.337 |  |
|  |  | C5-C6 | 0.346 ± 0.367 |  |
|  |  | C6-C7 | 0.557 ± 0.619 |  |
|  | Lumbar Spine | L1-L2 | 0.432 ± 0.369 |  |
|  |  | L2-L3 | 0.394 ± 0.403 |  |
|  |  | L3-L4 | 0.423 ± 0.390 |  |
|  |  | L4-L5 | 0.410 ± 0.372 |  |
|  |  | L5-S1 | 0.352 ± 0.376 |  |
| Calcium | Cervical Spine | C2-C3 | 90% |  |
|  |  | C3-C4 | 85% |  |
|  |  | C4-C5 | 85% |  |
|  |  | C5-C6 | 85% |  |
|  |  | C6-C7 | 86% |  |
|  | Lumbar Spine | L1-L2 | 84% |  |
|  |  | L2-L3 | 92% |  |
|  |  | L3-L4 | 88% |  |
|  |  | L4-L5 | 78% |  |
|  |  | L5-S1 | 82% |  |
| Vacuum disc | Cervical Spine | C2-C3 | 86% |  |
|  |  | C3-C4 | 90% |  |
|  |  | C4-C5 | 85% |  |
|  |  | C5-C6 | 78% |  |
|  |  | C6-C7 | 66% |  |
|  | Lumbar Spine | L1-L2 | 55% |  |
|  |  | L2-L3 | 65% |  |
|  |  | L3-L4 | 69% |  |
|  |  | L4-L5 | 63% |  |
|  |  | L5-S1 | 41% |  |
| 1c. Nitisinone Prediction |  |  |  |  |
|  | Region | Intervertebral Space | Test Accuracy |  |
|  | Cervical Spine | C2-C7 | 83% (total)<br>68% (“on treatment” accuracy) |  |
|  | Lumbar Spine | L1-S1 | 87% (total)<br>73% (“on treatment” accuracy) |  |

Figure 2a shows occlusion maps, along with the original images, for global score predictions. The cervical occlusion maps tend to focus on the central neck and spine, while lumbar images have more variable foci.

**Figure 2.**
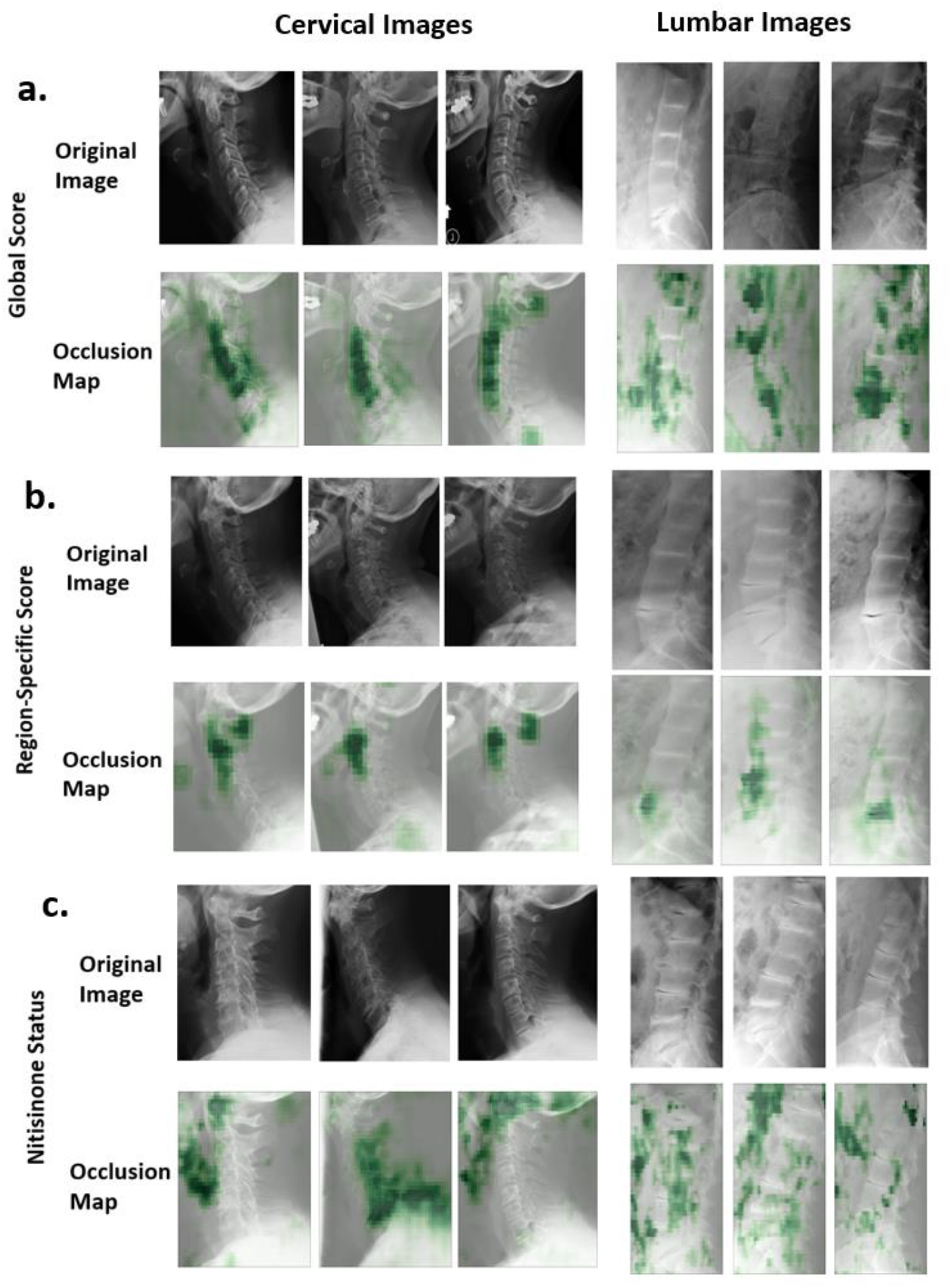
Examples of saliency maps for model prediction of (a) global scores, (b) region-specific scores, and (c) nitisinone status on cervical and lumbar AKU radiographs. Green, shaded regions indicate regions of model attention when predicting each metric for the entire cervical or lumbar image.

### Region-specific score

Table 1b presents narrowing, calcium, and vacuum disc estimates for individual cervical and lumbar IVS. For narrowing in the cervical IVS, the model obtained a range of 0.191-0.557 points from the expert scores. Predictions for the C6-C7 IVS showed the largest score differential from expert scores. In the lumbar spine, the model achieved a consistent range of scores across all IVS (0.352-0.432 points from the expert scores).

For calcium, cervical IVS predictions obtained high accuracy (85-90%), whereas lumbar IVS exhibited wider variability (78-92%). For vacuum disc, accuracy ranged from 66-90% in cervical IVS and was less consistent (41-69%) in lumbar IVS. The C6-C7 and L5-S1 IVS exhibited the lowest accuracy.

Figure 2b presents example occlusion maps and original images for region-specific predictions. Both the cervical and lumbar occlusion maps tend to focus on a single IVS.

### Nitisinone status

Table 1c summarizes nitisinone status predictions for cervical and lumbar images (39 images without treatment and 22 with treatment). For the cervical spine, the model achieved a total accuracy of 83%, with 68% accuracy for “on treatment” predictions. In the lumbar spine, performance was slightly higher, with a total accuracy of 87%, and 73% accuracy for “on treatment” predictions.

Figure 2c presents example occlusion maps and original images for nitisinone status predictions. As described (Discussion), these seem to focus on areas that may logically be affected by disease and treatment, including regions that involve ligaments, cartilage, and other connective tissue. Regarding potential confounders, there were no statistically significant differences across nitisinone status groups for age, sex, severity score, or time on treatment (Figure S3, Table S5).

Figure 3 shows the comparison of our model to expert clinicians at identifying nitisinone status. Our model (77% accuracy) outperformed human participants (51% accuracy) (p = 2.0 × 10^-9^) on a full test set of 61 images. When considering only the “on nitisinone” images in the surveys, the model (67%) outperformed human participants (33%) (p = 5.6 × 10^-8^).

**Figure 3.**
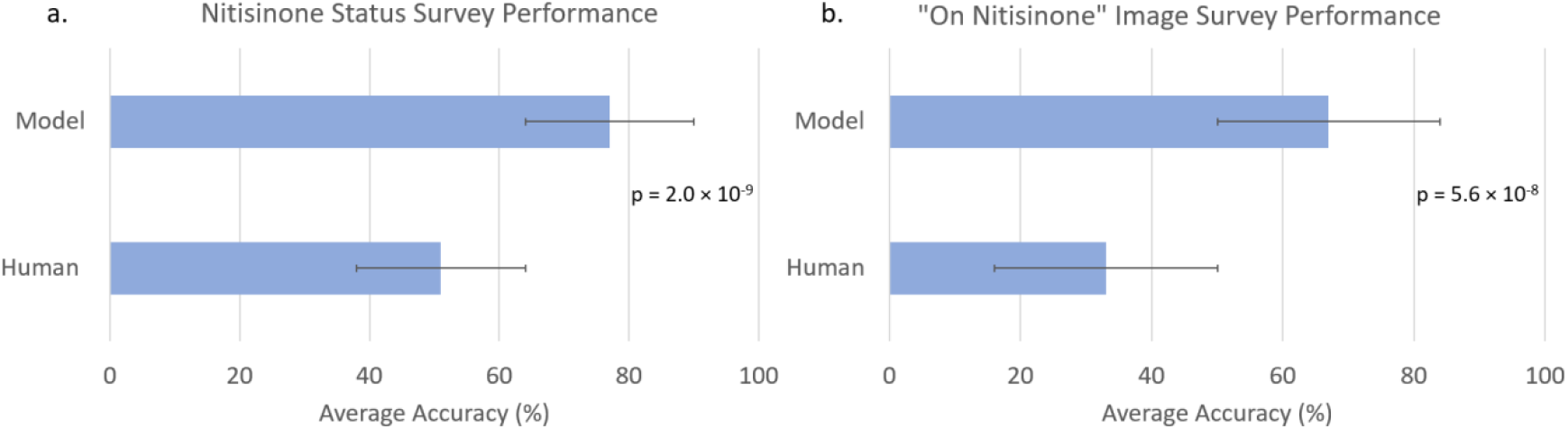
Comparison of nitisinone status detection performance between a DL model and human expert (geneticists and radiologists) on cervical and lumbar spine x-ray images for (a) all test images (Human = 51%, Model = 77%, n=61 images) and (b) “on nitisinone” only images (Human = 33%, Model = 67%, n=22). Average accuracy is shown for both the model and humans, with error bars representing standard deviation. The model outperforms human expert participants, achieving significantly higher accuracy in both the entire test set and the “on nitisinone” subset (p = 2.0 × 10^-9^, p = 5.6 × 10^-8^).

## Discussion

In this study, we trained DL classifiers to estimate severity scores of individuals with AKU based on cervical and lumbar radiographs.

As described (Supplementary material), we applied YOLO and SAM to segment each IVS and then trained EfficientNet on each segmented IVS.^1,20^ This yielded less accurate results than training the model using entire radiographs. This may be due to high correlations among IVS within a radiograph (Tables S2-S3). This is logical since AKU will typically affect multiple IVS.^21^ Thus, even when assessing a single IVS, other IVS may provide useful information. Further, other (non-IVS) areas may correlate with IVS disease; for example, there may be effects on other anatomical areas. Therefore, while we explored segmentation methods, our analyses ultimately focused on investigating a DL model to predict AKU spinal disease on full radiographs.

When considering the full radiograph, we first assessed the model’s ability to predict global severity, as this provides key information related to disease status.^21^ For global scores, the model achieved an average deviation of 1.72 ± 1.96 points from the ground truth cervical scores and 2.51 ± 1.96 points from the ground truth lumbar scores. Notably, these performance metrics are comparable to the minimal detectable change with 95% confidence interval (MDC_95_ ) value calculated by experts during manual annotation: 1.40 points for cervical scores and 1.61 points for lumbar scores, indicating that our models align closely with manual expert annotation. Additionally, linear regressions show strong correlations between the ground truth global severity score and the predicted global severity (R^2^ = 0.9441 for cervical and R^2^ = 0.8675 for lumbar, Table S4). Given the slow-progressing nature of AKU, the model reliably estimates overall severity within 1-2 years of disease progression (1.2-1.3 points = 1 year, per preliminary data from studies on this same dataset). In ongoing studies, experts reliably correlate the radiographic score with clinical assessments for AKU progression, such as pain levels, lumbar flexibility, or other physical functioning tests, establishing these scores as valuable clinical markers of disease progression over time. The region-specific models, while less useful in clinical practice, can help provide more granular estimation of AKU in terms of calcium, narrowing, and vacuum disc.

Expert clinicians look for a combination of different features (calcium, narrowing, and vacuum disc) in each IVS when assessing the clinical manifestations of AKU. Thus, we trained models to do the same (Table 1a, 1b). The models predicted narrowing with the highest accuracy (74-97%). Narrowing is very intuitive and can be easily observed as the physical distance between vertebrae. Additionally, narrowing was graded on a scale, so the expert clinicians could be specific about the amount of narrowing, providing more training information to the model. Narrowing performs with slightly higher accuracy for the cervical than the lumbar spine (0.338 versus 0.402 points differential from the ground truth, on average). Expert clinicians suggest this may be because lumbar spine IVS are larger than cervical IVS, making relative narrowing difficult to grade. Additionally, clinicians indicate that “normal” spines are easier to grade, and as lumbar spines are more affected, this may impact results (Figure S3).

Calcium was the next best predicted metric (78-92% accuracy) and also performs slightly better in the cervical spine. As with narrowing, this may correspond with the more severe lumbar findings. Lumbar regions are more obscured by other anatomical structures than cervical regions, and we noticed that the quality (e.g., contrast and clarity) of the lumbar images was more disparate than cervical images. Given our small dataset, it is thus expected that the cervical images perform slightly better due to their uniformity and lower severity.

Finally, vacuum disc underperforms and follows the trend of cervical versus lumbar accuracy. Vacuum disc was reported as more subjective for expert clinicians to grade, particularly in the lumbar spine. For example, clinicians described needing to adjust the contrast on radiographs to assess vacuum disc. Severe narrowing affects vacuum disc as well as calcium visibility, leading to more non-uniform cases of vacuum disc in the lumbar spine. This makes it difficult to build a training dataset that accurately represents the ways vacuum disc appears. Additionally, both calcium and vacuum disc can vary with respect to disease severity, narrowing status, and posture or angle of the images. However, due to subjectivity involved and non-linear nature of calcium and vacuum disc progression, our expert clinicians graded these as either absent or present. For example, both partial and total calcification would be classified as “present”, making model categorization potentially challenging.

In general, the regions close to the edges of the images (C6-C7, L5-S1) performed with lower accuracy. In our dataset, the outer areas of images tended to be less uniform and more obscured, often affected by overlapping anatomical structures (Figure S4). Therefore, clinicians were often unable to grade these regions; images without scores were removed from the dataset, reducing the sample size (Table S1).

Further, spine angulation can vary widely depending on a person’s posture, which can affect IVS appearance. In such cases, it can be hard to classify the images using small sample sizes.

To ensure the classifiers behave as expected, we generated and examined occlusion maps (Figure 2). In general, these maps showed that the model identified appropriate clinical patterns, rather than unrelated parts of the image (such as R or L metal markers).

While the global severity score may be a more clinically useful metric of disease progression in AKU, the region-specific analyses provide more context to help understand global scores. Together, the global and regional models may provide a more complete picture of AKU severity. Additionally, the relative importance of global versus regional severity may differ in other disease processes, such as conditions that manifest more focally; thus, it may be important for DL models to be able to consider both approaches.^22^

Our final analysis explored the ability to identify nitisinone status, which is particularly intriguing because clinical experts have not identified radiographic signs that could reliably indicate this. However, similar to findings in other studies—such as DL models predicting sex from retinal images—our results suggest that models may be able to detect patterns beyond human perception.^15^ The model achieved an accuracy of 83-87% on the full test set and 68-73% for individuals on nitisinone. We did not find evidence of logical confounders, but there may be factors that were not considered (Figure S3, Table S5).

A fascinating aspect of the nitisinone analysis involved occlusion maps. In cervical images, the model often highlighted the laryngeal region, ligaments, cartilage, and other connective tissue (Figure 2). In lumbar images, the focus was less consistent but included areas such as the aortic region, ligaments, and cartilage. These findings are potentially consistent with nitisinone affecting HGA deposition in connective tissue-rich regions, which could change the calcification of tissues in a subtle way detected by the model (but not humans). Although the saliency maps highlight regions such as the larynx, aortic region, and other connective tissues, none of the survey participants wrote comments about noticing these regions (see Supplementary material).^23^

Our study has limitations, including the small dataset. Additionally, the radiographs were collected over a 20-year period, during which variability in imaging techniques and equipment could yield inconsistencies. For example, lumbar images varied in terms of features like contrast, clarity, and resolution. This could pose challenges for the model in recognizing subtle features. Other limiting factors included the use of discretized scores by experts who manually graded the radiographs, as disease progression is continuous; the inherent subjectivity of human assessments suggests the benefit of standardized grading protocols or finer grading scales. Other limitations are discussed in the Supplementary material.

Despite these limitations, the study highlights the promise of DL in identifying clinically relevant patterns in rare diseases and small datasets and identifying features that may not otherwise be recognizable by the human eye.

## Data Availability

Data and code availability are described in the manuscript and can be found at: https://github.com/flahartyka/AKU-progression-efficientnet. We do not release the original dataset of AKU radiographs for patient privacy reasons and related to IRB requirements. If readers are interested in accessing the AKU dataset, they can contact the investigators of the clinical trial (00HG0141/NCT00005909), some of whom are co-authors on this study.

## Details of ethics approval

Applicable portions of the study have been approved by the following NIH IRB#: 00HG0141, 05HG0076, 000547, and 002349, and appropriate consent was obtained from all research participants in accordance with the approved protocols. These NIH IRB protocols correspond to the original 20-year clinical trial of individuals with AKU (00HG0141), the clinical trial of individuals on nitisinone treatment (05HG0076), the analyses done in this study including using deep learning to analyze these images (000547), and the survey exemption protocol to send out surveys of these images to clinical geneticists and radiologists (002349).

