## Supplementary Material for "Deep Learning Study of Alkaptonuria Spinal Disease Assesses Global and Regional Severity and Detects Occult Treatment Status"

**Supplementary Methods**

*Preparing the Dataset and Scoring X-rays*

Data were obtained through National Institutes of Health (NIH) Protocols related to the study of Alkaptonuria (AKU) (IRB# 00HG0141 and 05HG0076), and all research participants provided informed consent; additional analyses done in this study were permitted via NIH IRB# 000547. As part of these studies, X-ray images were taken of the cervical and lumbar spine of patients with AKU; radiographs (including multiple images from the same individual) were obtained at the NIH Clinical Center over a 20-year period (January 2003 to May 2023). A total of 409 sets of radiographs were taken, and we divided the data into 338 training images and 71 testing images. The training and testing set contain unique patients, thus avoiding data-leakage where images of the same patient are included in both sets. After removing all the poor-quality images, we analyzed 397 cervical spine and 395 lumbar spine images in total (an average of 3 cervical images per patient, and likewise for lumbar spine). Statistics about age, sex, spinal disease severity of patients the AKU dataset as well as the number of images which were manually gradable for each region are shown in Table S1.

Per previous and ongoing research methods of assessing in the spinal effects of AKU, disease severity was evaluated based on three primary metrics: narrowing, calcium, and vacuum disc, which were chosen based on previous research and were specifically selected to match ongoing NIH research assessing AKU progression.[1] Each cervical and lumbar lateral X-ray image was manually graded by experts using a standardized schema currently used by collaborating researchers. In summary, experts graded each intervertebral space (IVS) in the cervical spine from C2 to C7 and each IVS in the thoracolumbar spine from T10 to S1. For each segment, IVS were scored based on three criteria: joint space narrowing, intervertebral disc calcification, and vacuum disc phenomenon. Narrowing was graded as absent (no narrowing, score = 0), mild (less than 1/3 of the disc space narrow, score = 1), moderate (1/3 to 2/3 of the disc space narrow, score = 2), severe (more than 2/3 of the disc space narrow, score = 3), or fused (no disc space, score = 6). Calcium and vacuum disc were scored as either absent (score = 0) or present (score = 1).[2] If the narrowing was categorized as fused, calcium and vacuum disc presence were not gradable but were still accounted for by the higher scoring for fused (hence, the maximum score of disc space narrowing is 6 instead of 4).

In this study, however, we excluded the thoracic regions (T10-T11, T11-T12, and T12-L1). These regions were originally included in the expert clinician's manual grading, where they calculated the thoracolumbar score of 48 total points. We decided to exclude the 3 thoracic IVS and solely focus on the cervical and lumbar spines for our DL models (total score of 30 points for both cervical and lumbar spines). This was done because, based on clinical experts' advice, the thoracic regions were often obscured (e.g., by overlapping anatomy) or unavailable on lumbar X-ray images, requiring separate thoracic images. Therefore, after dataset preparation, our dataset for the thoracic region was much smaller than our datasets for the cervical and lumbar regions, and we were unable to create a well-balanced test set for all three regions. Our study utilized this manually graded AKU dataset to train and evaluate several image classifier approaches designed to predict these metrics.

X-ray images were cropped to remove the black border surrounding each image, zoomed, and centered using an automated cropping method. Finally, for standardization, all images were oriented to face the same direction. R and L metal markers and other markings on the X-rays, when present, were not intentionally removed, but we noted that these markings occur randomly (and did not appear to be areas of focus when occlusion maps were assessed). Thus, they are not likely to provide significant unintended signal. The images produced via these initial processing steps are referred to as the "full images".

Tables S2 and S3 show the correlation matrices of severity scores (calcium, narrowing, and vacuum disc scores combined) for all intervertebral spaces (IVS) in both the cervical and lumbar spines. Due to strong positive correlations for each IVS, it is likely that the severity of each region can provide information that allows accurate prediction of the severity of other regions. That is, the findings in one IVS tend to correlate with the findings in another IVS. Thus, we trained our image classifier on full images so that information from all regions could be included in the images.

Originally, we wanted to crop as close to spinal region as possible; thus, removing areas like the base of skull, larynx, back of neck. However, these other signals (although they do not overlap with the IVS) could also be important for some assessments, such as those described in the Discussion regarding the nitisinone experiment. We also used segmentation methods to further process the full images into spine-only images and individual vertebral images. This allowed additional analyses on individual portions of the images. Descriptions and examples of segmentation methods, further image processing, and evaluation of the segmented images can be found in the Supplementary material below. Next, we selected an image classifier model and evaluated our full images through a variety of experiments.

*Model Selection and Image Evaluation*

We chose EfficientNet because it performs well using relatively few parameters.[3] Moreover, Efficient-Net has been finetuned successfully for other small datasets related to the study of rare conditions.[4-6] To train EfficientNet, we used 2 v100x GPU and batch size 64. We used Adam optimizer with learning rate 0.00003. We loaded the pre-trained weights of ImageNet, and then finetuned these weights using our AKU X-ray images.[7] To improve model accuracy and sample size, 400 images from publicly available cervical and lumbar spine radiographic datasets (CSXA and BUU-LSPINE) were incorporated into our AKU dataset and labeled as “normal”. [8,9] This was done to increase the total training and testing dataset sizes because the original datasets of 397 cervical and 395 lumbar AKU images performed with slightly better accuracy across all experiments when these additional 400 "normal" images were added. All models were trained with 5-fold cross validation. Thus, there are 5 models, and for a test image, the final prediction is the average outcomes of these 5 models. Our trained models and code to reproduce the results are available at: https://github.com/flahartyka/AKU-progression-efficientnet. We acknowledge that there are models that may outperform EfficientNet at these tasks; however, in this paper, as a proof of concept, we aimed to evaluate whether a DL classifier can obtain reasonable results on our dataset. Further study could include exploring how other image classifiers and ensemble approaches would improve model performance.

Although images were taken from the same patient at different time points, we treated these images as if they were independent. Thus, the model was trained to estimate disease severity at just a single time point. We noted that the model still produces consistent predictions when considering these images at single time points; for example, one patient in the test set has a global score prediction of 11 at time point 1, and at time point 2 (three years later) has a global score prediction of 15. We chose to do this since the manual expert scoring system we used to analyze our results also treated the images independently, and as manifestations may change unpredictably over time, both in terms of how different regions of the spine are affected, and how different individuals are affected. Future work could include evaluating other DL image models for time-series datasets.[10]

Among different patients, the IVS can differ simply due to factors like age and sex or other individual characteristics of a person's spine. Thus, human annotators assessed the entire spinal region before determining the severity of an individual IVS. For this reason, we applied EfficientNet to the entire x-ray image when estimating the global and region-specific severity scores. As detailed below, we also implemented a segmentation-based approach for thoroughness. However, this method did not perform as well compared to evaluating all IVS simultaneously using the full image.

*Segmentation*

Segmentation methods were explored to automate ways of isolating and analyzing specific vertebrae and IVS. This was done with the hypothesis that removal of any random or unrelated signal within the X-ray images (such as letters or other parts of the X-ray) would allow the vertebral regions to be better isolated to evaluate severity. Two methods of segmentation were used to isolate the cervical and lumbar vertebrae into spine-only images and individual vertebral images: Finetune Segment Anything Model (finetune-SAM) and YOLO.[11,12]

Spine-only images exclude extraneous areas and areas that may initially seem irrelevant such as the larynx and the skull. Individual vertebral images include only the region that includes a single vertebral body. Both finetune-SAM and YOLO models were finetuned on publicly available CSXA (cervical) and BUU-LSPINE (lumbar spine), with ~5000 images in each.[8,9] These datasets provided the coordinates for each individual vertebral body such that masks outlining the C2-C7 regions of the cervical spine and L1-S1 regions of the lumbar spine could be generated for each image (Figure S1). Then, both models were trained on these publicly available images and their corresponding masks.

Finetune-SAM is a model based on the Meta Segment Anything Model (SAM); it allows for finetuning SAM on small datasets.[11] Following Gu et. al., a single GPU was used to finetune the base SAM on CSXA and BUU-LSPINE datasets, respectively.[8,9,11] No additional finetuning was done on our AKU images, and the final inference step was done based on transfer learning. Thus, we simply applied the trained models on our AKU dataset to produce masks of IVS, individual vertebral bodies, and the spine-only region, such as those shown in Figure S2.

Although finetune-SAM is based on state-of-the-art SAM, at the time of our paper, finetune-SAM code did not have the functionality to label specific vertebral bodies. To address this, the YOLO model was employed to predict both the segmentation and labeling of individual IVS, enabling further parsing of images into specific regions. The YOLO implementation from Ultralytics uses bounding boxes instead of the natural contour of the objects.[12] Thus, we converted the human annotations of the vertebral bodies from the 2 available datasets into bounding boxes. After finetuned on CSXA and BUU-LSPINE, YOLO was then applied on our AKU dataset to generate labeled masks for each vertebra (without any additional training on our AKU dataset).

Four types of segmentation were applied to the cervical and lumbar images, summarized in Figure S2. Subsequently, YOLO-labeled AKU segmented areas-of-interest were analyzed using EfficientNet to predict narrowing and calcium. Since we applied transfer learning without finetuning on AKU images, only 315 of 400 cervical images and 300 of 400 lumbar images were successfully segmented by the YOLO model. While segmentation facilitated the division of full images into individual region-specific areas, these segmented areas yielded accuracy below 50%, and correlation matrices for cervical and lumbar spines indicated that analyzing full images was more effective (see Discussion in main paper). Consequently, our subsequent analyses focused on the full images (see details in main manuscript sections).

*Nitisinone Status Survey Setup*

As previously noted, nitisinone status is not typically discernible to experts analyzing these X-rays, as no known marker for nitisinone treatment presents in this type of imaging. Biochemical geneticists and radiologists were recruited for this study. To identify participants, we obtained names of known alkaptonuria (or related conditions) expert geneticists as well as radiologists, through professional networks and publications in the field.[5,6,13,14] Participants were recruited via email and provided a link to 1 of 8 possible surveys, which only varied based on the images shown (the surveys showed different images to ensure that all test images could be evaluated by these participants). Both the email recruitment document, as well as the survey directions requested that only physicians with an expertise in alkaptonuria participate in the survey. Of the 64 international medical geneticists and radiologists solicited, 12 completed the surveys (9 medical geneticists and 3 radiologists). The survey portion of the study was approved as IRB exempt by the NIH IRB (IRB# 002349).

*Comparison of Physicians' Classification Accuracy with Model Predictions*

We assessed alkaptonuria expert medical geneticists' and radiologists' ability to recognize spinal X-ray images of individuals undergoing nitisinone therapy via surveys sent using Qualtrics (Provo, Utah, United States). Eight versions of the survey were generated to provide a reasonable number of images to classify (n=20) while allowing for all images in the test set to be evaluated (n=61). Each survey included 10 cervical spinal X-rays (5 from patients receiving nitisinone therapy and 5 not receiving therapy) and 10 lumbar spinal X-rays (5 from patients receiving nitisinone therapy and 5 not receiving therapy). Participants were not told prior to the survey the numbers of images of each type that they would encounter. Participants were shown each of the 20 images one at a time and asked to classify the image (whether it showed a patient receiving nitisinone therapy or not), and were provided the option to enter text explaining their reasoning for classification. At the end of the survey, participants were asked to rate their confidence level for identifying nitisinone status in spinal x-rays on a scale of 0 (not at all confident) to 100 (highly confident). Participants also answered demographic questions (medical specialty and years of experience). Surveys and full responses can be found at: https://github.com/flahartyka/AKU-progression-efficientnet. Accuracies were calculated by comparing the participant's prediction and the model's prediction to the ground truth nitisinone status of the patient. Then, the total number of images correct over the total number of images in the survey (n=20) was calculated. The average accuracy was taken of all surveys completed and compared to the average accuracy of the model. T-test was performed to assess the statistical difference between the model and the expert cohort.

**Supplementary Figures**

**Figure S1. Example cervical image and segmentation masks used to train finetune-SAM and YOLO models.** Three types of masks were generated of image 0008035 from CSXA dataset: intervertebral space masks, vertebral body masks, and spine-only masks. Images and masks created from CSXA and BUU-LSPINE datasets.[8,9]

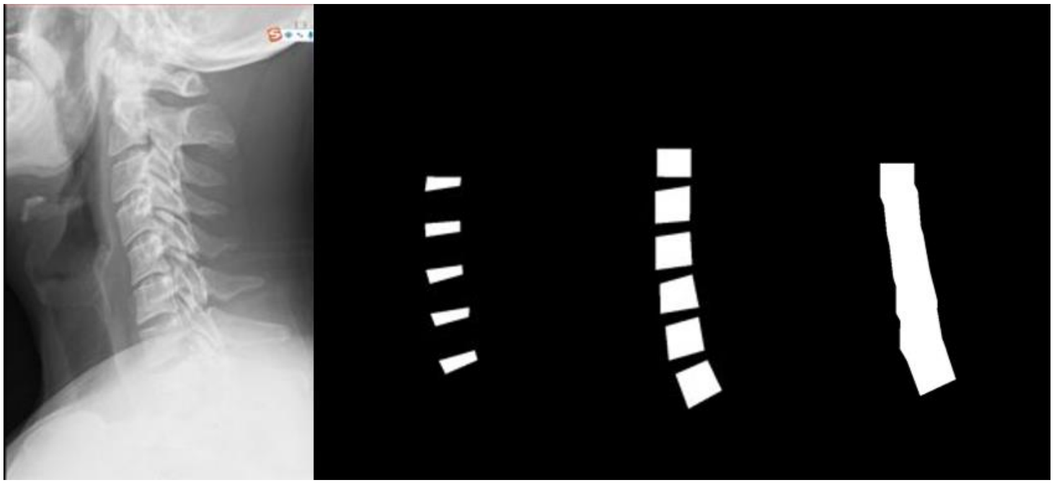

**Figure S2. Segmentation and labeling of cervical and lumbar spine Alkaptonuria X-ray images.** (a) Original cervical and lumbar spine X-ray image. (b) Segmentation of cervical and lumbar vertebral bodies. (c) Segmentation of cervical and lumbar intervertebral spaces. (d) Segmentation of the entire cervical and lumbar spine region including both the vertebral bodies and IVS. (e) YOLO-based segmentation with labeling of individual vertebral body.[8,9]

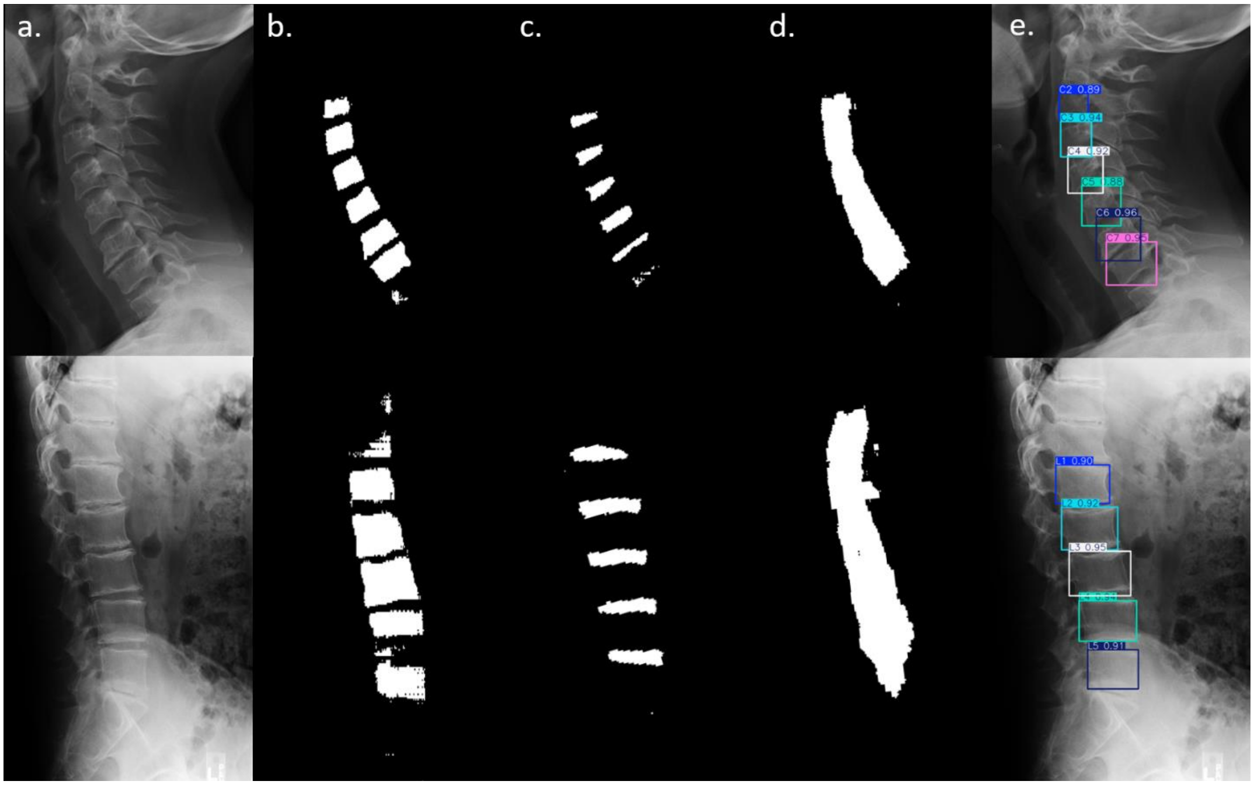

**Figure S3. Distributions of (a) cervical severity score (b) lumbar severity score and (c) age according to ground truth nitisinone status.** These three metrics were assessed as potential confounders for EfficientNet nitisinone status prediction. No group comparisons show statistically significant differences.

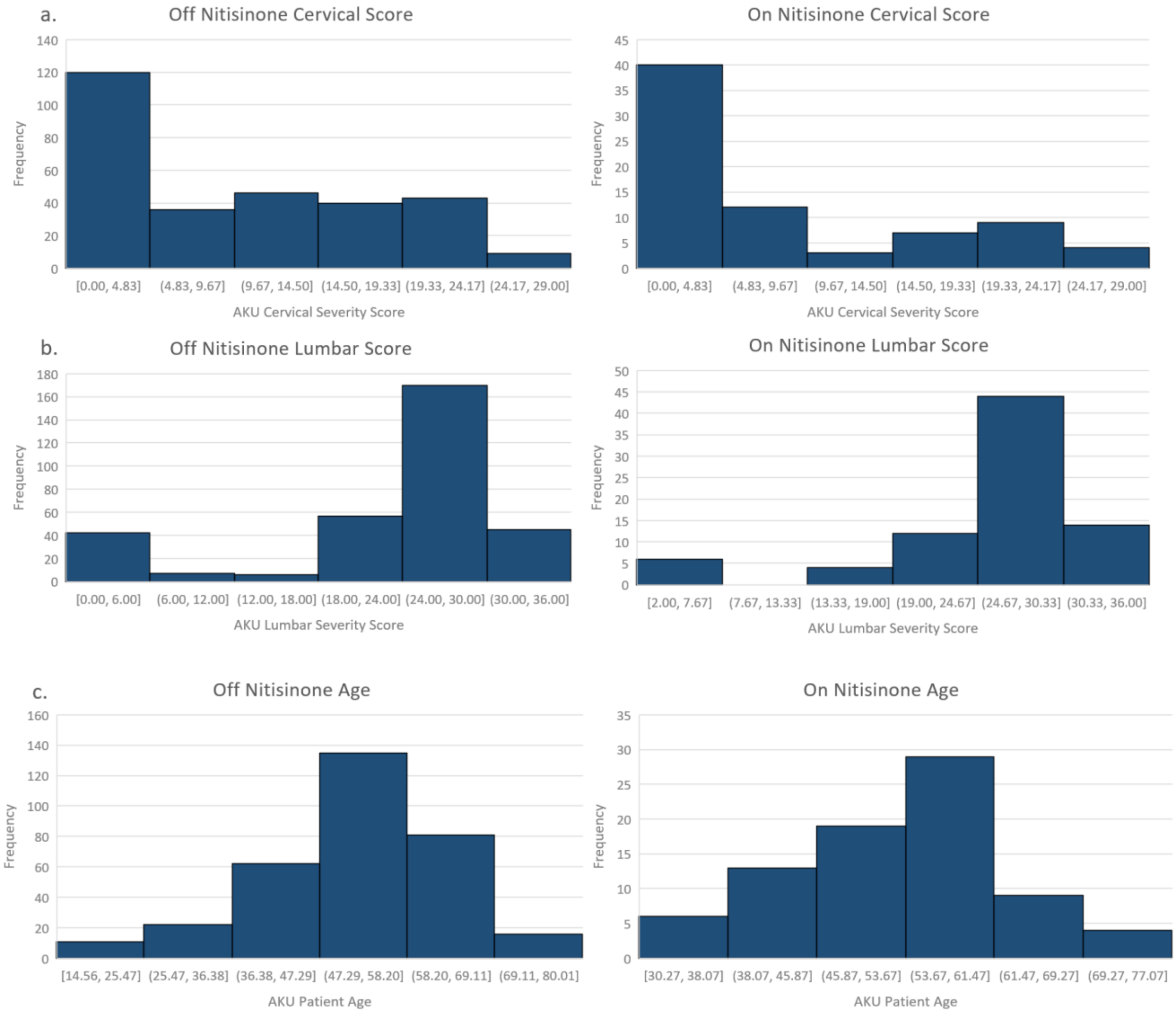

**Figure S4. Examples of Alkaptonuria X-ray images where the most distal intervertebral spaces (IVS) (C6-C7 and L5-S1) are visible and obscured.** These examples show the variability of these outer IVS and where these regions may be obscured by other bones and internal structures within the frame.

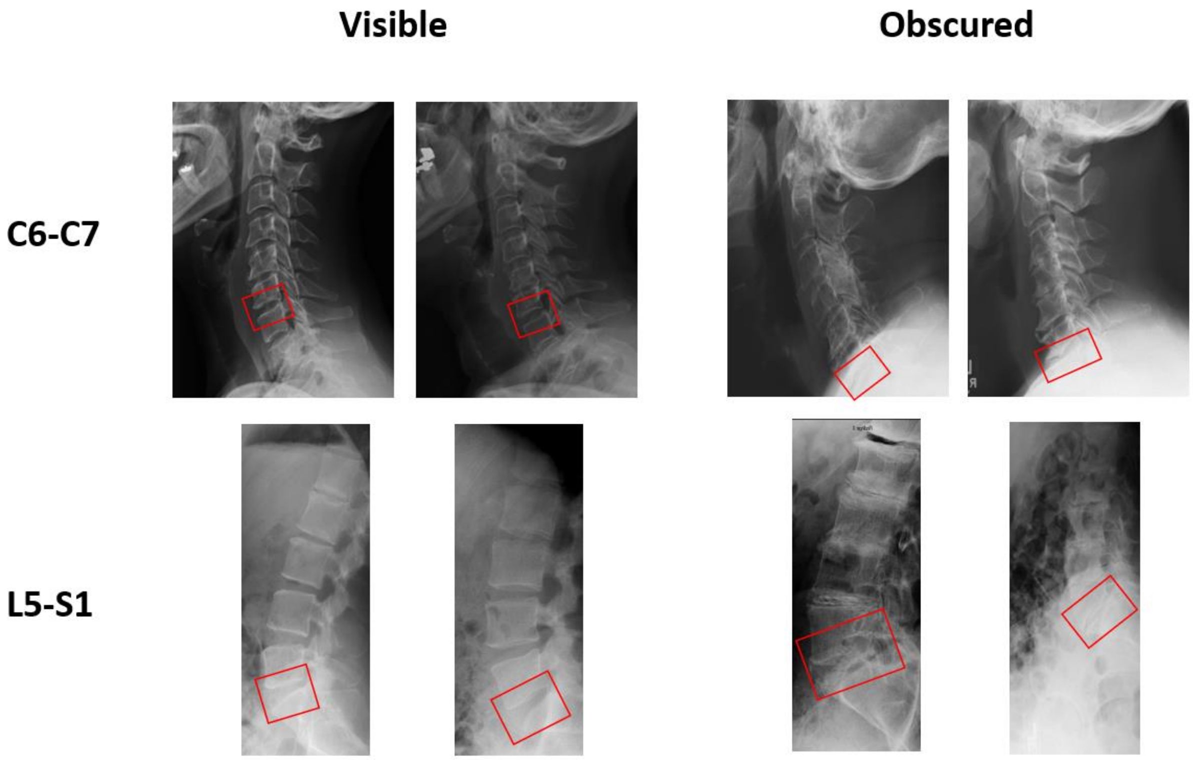

**Tables**

**Table S1. (a) Alkaptonuria dataset statistics and (b) numbers of images with scores for each region after pre-processing steps.** Ratios and specific numbers of images associated with age, sex, nitisinone status, severity scores, training/testing cohorts.

| **1a. Dataset Statistics** | |
|---|---|
| **Age range (years)** | 14.5-80 |
| **Mean age (years)** | 52.3 |
| **Male: Female ratio** | 0.60: 0.40 (244: 165) |
| **On: Off Nitisinone ratio** | 0.20: 0.80 (80: 329) |
| **Cervical Mild:Moderate:Severe ratio** | 0.63: 0.24 :0.13 (250: 95: 52) |
| **Lumbar Mild:Moderate:Severe ratio** | 0.13: 0.063: 0.81 (51: 25: 319) |
| **Training: Testing ratio** | 0.83: 0.17 (338: 71) |
| **1b. Numbers of Images** | |
| **Total sets** | 409 |
| **Complete scores (Cervical, Lumbar)** | 370, 398 |
| **Total after removing bad quality or poorly cropped images (Cervical, Lumbar)** | 397, 395 |
| **Complete scores after removing poor quality or poorly cropped images (Cervical, Lumbar)** | 370, 393 |
| **C2-C3, C3-C4, C4-C5, C5-C6, C6-C7** | 397, 396, 397, 393, 371 |
| **L1-L2, L2-L3, L3-L4, L4-L5, L5-S1** | 395, 395, 394, 393, 394 |

**Table S2. Correlation matrix for total severity scores at cervical spine intervertebral spaces.** Pearson correlation coefficients for total severity scores between intervertebral spaces in the cervical spine (C2-C3 through C6-C7). Higher values indicate stronger positive correlations (max = 1).

| | C2-C3 | C3-C4 | C4-C5 | C5-C6 | C6-C7 |
|---|---|---|---|---|---|
| **C2-C3** | 1 | | | | |
| **C3-C4** | 0.695785 | 1 | | | |
| **C4-C5** | 0.679339 | 0.77668 | 1 | | |
| **C5-C6** | 0.655277 | 0.672771 | 0.712242 | 1 | |
| **C6-C7** | 0.582196 | 0.607959 | 0.655511 | 0.78109 | 1 |

**Table S3. Correlation matrix for total severity scores at lumbar spine intervertebral spaces (IVS).** Pearson correlation coefficients for total severity scores between IVS in the lumbar spine (L1-L2 through L5-S1). Higher values indicate stronger positive correlations (max = 1).

| | L1-L2 | L2-L3 | L3-L4 | L4-L5 | L5-S1 |
|---|---|---|---|---|---|
| **L1-L2** | 1 | | | | |
| **L2-L3** | 0.751277 | 1 | | | |
| **L3-L4** | 0.721015 | 0.819199 | 1 | | |
| **L4-L5** | 0.705676 | 0.754729 | 0.81987 | 1 | |
| **L5-S1** | 0.737322 | 0.717984 | 0.7621 | 0.769688 | 1 |

**Table S4. Cervical and lumbar global score regression results between the ground truth severity score to the predicted severity score.** $R^2 = 0.9441$ for cervical scores and $R^2 = 0.8675$ for lumbar scores, indicating high agreement between the ground truth and EfficientNet predicted global severity scores. $R^2$ was computed via a linear regression line without the intercept.

| Region | Comparison | $R^2$ |
|---|---|---|
| **Cervical Spine** | Ground truth global score versus predicted global score | 0.9441 |
| **Lumbar Spine** | Ground truth global score versus predicted global score | 0.8675 |

**Table S5. Assessment of potential confounders for EfficientNet nitisinone status prediction.** T-tests assuming equal variances were conducted to compare the on treatment versus off treatment group (with the exception of the "time on treatment" metric) for confounders such as severity score, age, and sex. No group comparisons showed statistically significant differences.

| Confounder | Comparison (Group 1 versus Group 2) | Group 1 Mean | Group 2 Mean | p-value |
|---|---|---|---|---|
| **Cervical Severity Score** | On treatment versus off treatment | 8.0 | 9.5 | 0.173567598 |
| **Lumbar Severity Score** | On treatment versus off treatment | 25.3 | 23.3 | 0.081956408 |
| **Age (years)** | On treatment versus off treatment | 52.6 | 52.1 | 0.712231068 |
| **Sex** | On treatment versus off treatment | 0.43 (43% female, 57% male) | 0.40 (40% female, 60% male) | 0.69132472 |
| **Time on Treatment (years)** | Model correct versus incorrect | 2.99 | 1.96 | 0.323274539 |

**Table S6. Accuracy of C3-C4 calcium level for cervical X-rays before and after adding 400 images from the CSXA dataset.** Calcium statuses are categorized as present, absent, or fused, and reported as the accuracy of the model in making the correct prediction. Accuracy improves when training with 400 additional free images rather than without the free images.

| AKU-Only Dataset | AKU Dataset + 400 CSXA Images |
|---|---|
| 81% | 85% |
